# Texture-based Probability Mapping in cardiac LGE Images: A novel automatic assessment of myocardial injury following revascularized STEMI

**DOI:** 10.1101/2023.09.20.23295871

**Authors:** Vidar Frøysa, Gøran J. Berg, Erlend Singsaas, Trygve Eftestøl, Leik Woie, Stein Ørn

## Abstract

**Background:** Scar size is critical to left ventricular (LV) remodeling and adverse outcomes following myocardial infarction (MI). Late Gadolinium-enhancement (LGE) in cardiac magnetic resonance imaging is the gold standard for assessing MI size. Texture-based probability mapping (TPM) is a novel machine learning-based analysis of LGE images. This proof-of-concept study investigates the potential clinical implications of temporal changes in TPM during the first year following an acute revascularized MI.

**Methods:** 41 patients with first-time acute ST-elevation MI were included in this study. All patients had a single-vessel disease and were successfully revascularized by primary percutaneous coronary intervention. LGE images were obtained two days, one week, two months, and one year post-MI. MI size by TPM was compared with manual LGE-based MI calculation, LV remodeling, and biomarkers.

**Results:** TPM showed a significant increase in infarct size from the second month through the first year (p<0.01). MI size estimated by TPM at all different time points demonstrated strong correlations with peak Troponin T levels. At one week, TPM assessment correlated positively with maximum C-reactive protein (r=0.54, p<0.01), and at two months, TPM positively correlated with N-Terminal Pro Brain Natriuretic Peptide.

**Conclusion:** This proof-of-concept study suggests that TPM may provide additional information to conventional LGE-based MI analysis of scar formation, LV remodeling, and biomarkers following an acute revascularized MI.

**Highlights:**

- Texture-based probability mapping (TPM) was used to analyze consecutive cardiac magnetic resonance images acquired during the first year after ST-elevation myocardial infarction (STEMI).
- TPM size was related to biomarkers of inflammation, myocardial injury and stress.
- TPM is a step toward automatic image processing.

## 1. Introduction

The size of the myocardial infarct (MI) is a crucial factor in the development of left ventricular (LV) remodeling and heart failure (HF) with subsequent increased morbidity and mortality ^1–7^. Assessment by late gadolinium-enhanced cardiac magnetic resonance imaging (LGE-CMR) is considered the gold standard for the in vivo assessment of MI size and scar development ^8, 9^. Current methods to determine MI size from LGE-CMR images are based on regional differences in pixel signal intensities ^10^.

The texture-based probability mapping (TPM) is a novel analytic method for assessing scar size in LGE-CMR images. The TPM method uses machine learning and a reference dictionary to analyze the texture of pixel matrices from LGE-CMR images to calculate MI size^11^. The TPM method can assess scar size in individuals with > one year old MI ^12^. Compared with traditional LGE-CMR analysis, the TPM method provides a potentially broader assessment of myocardial injury in patients with established MI, allowing assessment of both the infarcted and non-infarcted myocardium.

Since the TPM assessment is based on a reference of healed MI, it is unclear how it relates to the complex dynamic myocardial alterations following acute MI. This study, therefore, examines the temporal changes in TPM assessment during the first year after an acute MI using serial LGE-CMR images, comparing TPM with traditional LGE-CMR MI estimates, LV remodeling parameters, and biomarkers.

## 2. Material and methods

### 2.1 Study population

The present study analyzed serial digital LGE-CMR images from a unique group of patients admitted to the hospital due to first-time acute STEMI. All patients had occluded single-vessel disease with a culprit lesion in a large coronary artery’s proximal or middle part. All patients were treated successfully by primary percutaneous coronary intervention (PCI) without residual stenosis. Patients with evidence of prior MI or reinfarction during the first week after PCI were excluded from the study. The patients received anti-remodeling therapy according to conventional guidelines during the study. The study was approved by the Regional Ethics Committee at the University of Bergen and was conducted according to the Declaration of Helsinki principles. All patients gave informed consent before their inclusion in the study.

### 2.2 Cardiac Magnetic Resonance Imaging Exams

LGE-CMR was used to assess the patients at four preallocated time intervals after MI: two days, seven days, two months, and one year following PCI. CMR was performed with a 1.5 T Philips Intera R 8.3. All images were ECG gated and obtained during breath holding. Functional assessment of LVEF and LVEDV was performed according to current recommended standards using a steady-state, free precession sequence covering the entire left ventricle with 8-mm thick short-axis slices and inter-slice gaps of 2 mm. LV volumes were assessed on complete short-axis datasets in a random, blinded fashion using the View Forum^TM^ Software (Philips Medical Systems, Best, The Netherlands). Indices for LV volumes were obtained by correcting for body surface (LVEDVi).

Following the functional assessment, a gadolinium-based contrast agent, Omniscan®, was administered intravenously at a dosage of 0.25 mmol/kg (to reduce problems with early washout). LGE-CMR images were acquired 10-15 minutes later; using an inversion-recovery-prepared T1 weighted gradient-echo sequence with a typical pixel size of 0.82x0.82 mm^2^, without parallel imaging, covering the whole ventricle with short-axis slices of 10 mm thickness, without inter-slice gaps. Inversion recovery sequences were collected with individually adapted inversion times of 200 to 300 milliseconds to nullify normal myocardium. MI size was quantified based on these images by two experienced, independent, and blinded observers and is expressed as a percentage of total LV mass. A previous study describes The CMR protocol in detail ^5^.

### 2.3 Texture-based Probability Mapping of Cardiac Magnetic Resonance Images

The original LGE-CMR short-axis images were segmented with manual endo- and epicardium tracking to define the myocardium pixels by two experienced researchers (Fig. 2). Texture measure was calculated for each myocardial pixel following the manual segmentation. This texture measure corresponds to the representation error when matrices of grey-level intensities for every pixel’s spatial neighbourhood are represented by sparse linear combinations of atoms from two dictionaries. A classifier distinguishes between pixels from scarred and normal myocardial tissue. The classifier has been developed from the textural measure assessed in LGE-CMR images of patients with old myocardial infarction. Due to the high risk of ventricular dysrhythmia, these patients were treated with an ICD. The classifier utilizes Bayes’ decision theory, which includes calculating the probability density functions for the texture features extracted from scar and healthy myocardium. The classifier also calculates a prior probability function reflecting the mean value of the scar to normal myocardial size ratio in the training data set. The results of the TPM analysis can be visualized in different ways, like color coding the pixels according to their probability of being a scar.

One example of this visualization is a probability map (Fig. 1). Detailed descriptions of the TPM method and the classifier training have been provided in ref.^11, 13^. The TPM was computed in MATLAB (MathWorks, Natick, Massachusetts, USA). Some LGE-CMR exam images had 256 x 256 pixels resolution, and these were up-sampled before the import to MATLAB so that all images had 512 x 512 pixels resolution at the segmentation and TPM analysis. Images with original 512 x 512-pixel resolution were down-sampled to 256 x 256 and then up-sampled to their original pixel resolution to let all images go through the same process before analysis.

**Fig. 1.**
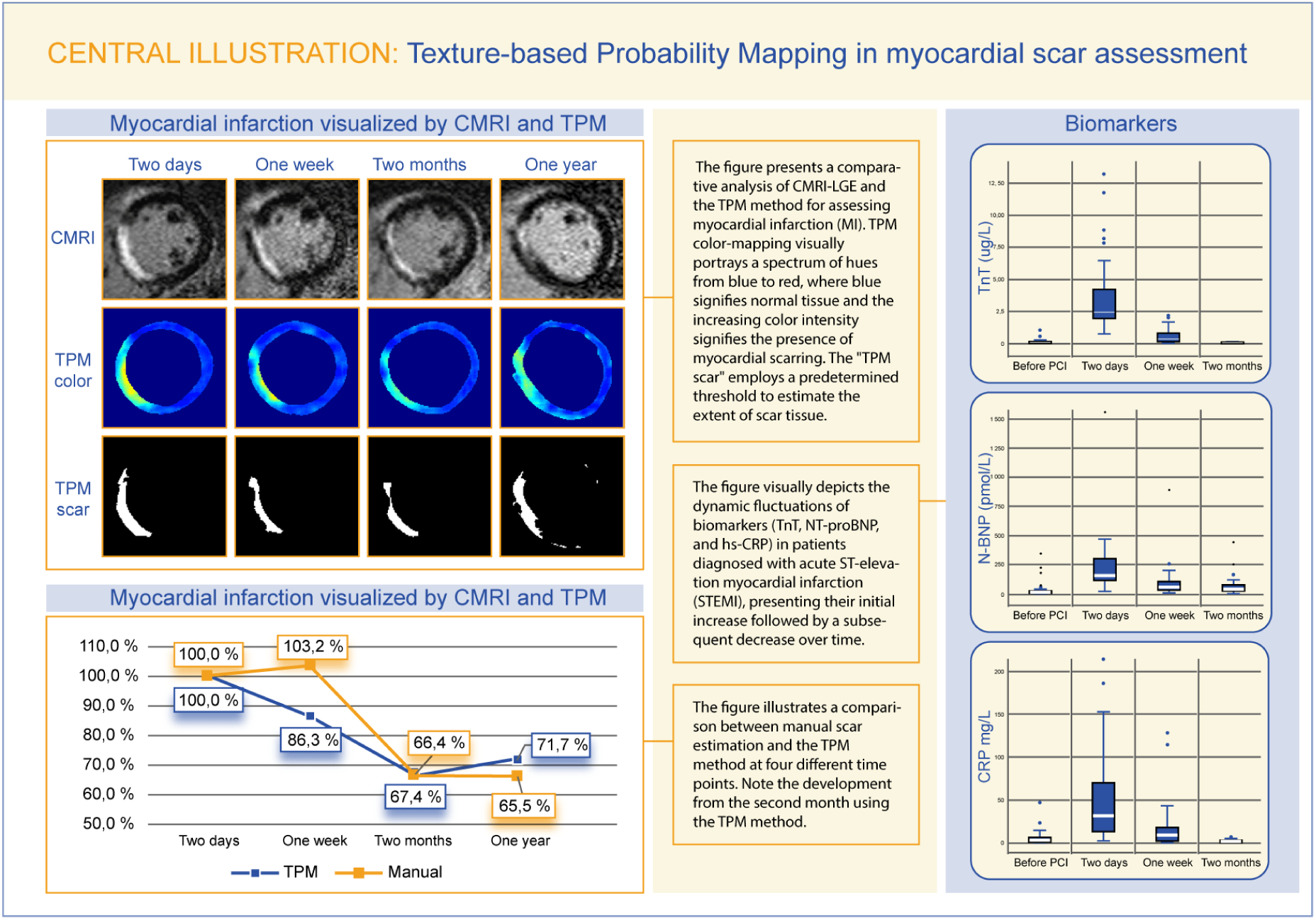
Central illustration.

The TPM analysis defines a cardiac segment as a myocardial region with pixel probabilities within a specified range.^13^

MVO within the infarcted myocardium in the acute phase of MI is a marker of profound ischemic damage to the myocardium^5^. This phenomenon is seen as black regions within the hyper-enhanced regions due to the obliteration of the microvasculature. The TPM method accounts for this through an algorithm to prevent MVO from being classified as normal myocardium^14^.

### 2.4 Biomarkers and Laboratory Analysis

Venous blood was collected from the patients before, two days, one week, and two months after PCI. To obtain platelet-poor plasma, pyrogen-free blood collection tubes were immediately immersed in melting ice (EDTA-containing tubes) and centrifuged within 20 minutes at 2500 g for 20 minutes. For serum, tubes without additives were kept at room temperature and centrifuged at 1000 g for 10 minutes after coagulation. The samples were stored at −80 °C. High-sensitive C-reactive protein (hs-CRP) concentrations were assessed by a particle-enhanced immunoturbidimetric method through the use of Roche ModularP automated clinical chemistry analyzer (Roche Diagnostics, Basel, Switzerland) and reagents of Tina-quant CRP (latex) assay (Roche Diagnostics). TnT concentrations were measured on Roche Elecsys 2010 (Roche Diagnostics), with the immunoassay TnT (Roche Diagnostics), using a biotinylated monoclonal TnT-specific antibody and a monoclonal TnT-specific antibody.

### 2.5 Statistical analysis

The statistical analysis was computed using a commercially available statistical calculation program (IBM SPSS version 21, IBM, Armonk, New York, USA). Continuous data with a normal distribution are reported as mean with a 95% confidence interval, and Non-normally distributed variables were presented as median with 25^th^ and 75^th^ percentiles. To compare related continuous data, the Wilcoxon signed ranks test was utilized. The Spearman method was used to analyze correlations. All p-values are two-tailed, and a p-value of less than or equal to 0.05 was considered statistically significant.

## 3. Results

### 3.1 Baseline characteristics

This study used serial LGE-CMR images from 46 patients in a previous single-center study^5^. Of these 46 patients, five patients were excluded due to reinfarction (n=2), withdrawal from the study (n=2), or incomplete data set (n=1). The remaining 41 patients (81% male with a mean age of 58 years) served as the study sample (Table 1).

**Table 1.** Baseline characteristics for all subjects (n=41)

|  |  |
| --- | --- |
| Age (years) | 58.3 ± 12.2 |
| Male gender | 33 (81%) |
| BSA | 2.0 ± 0.2 |
| Creatinine (μmol/L) | 71 (63, 96) |
| eGFR (mL/min/1.73m<sup>2</sup>) | 103 (80, 113) |
| Medical history |  |
| Hypertension | 10 (24%) |
| Hyperlipidemia | 10 (24%) |
| Smoking | 19 (46%) |
| Diabetes mellitus | 3 (7%) |
| Stroke | 1 (2%) |
| PVD | 2 (5%) |
| MI after the index event | 0 |
| Revascularisation after the index event | 0 |
| Drug therapy |  |
| Acetylsalicylic acid | 41 (100%) |
| Statin | 40 (98%) |
| Betablocker | 20 (49%) |
| ACE-I / ARB | 29 (71%) |
| Diuretic | 4 (10%) |
| Warfarin | 2 (5%) |
| Aldosterone antagonist | 7 (17%) |

Non-normal distributed continuous variables are expressed as median (25<sup>th</sup>, 75<sup>th</sup> percentile), and normally distributed data as mean ± SD. Categorical as absolute number (%). BP = blood pressure, MI = myocardial infarction, PVD = peripheral vascular disease, ACE-I = angiotensin-converting enzyme inhibitor, ARB = angiotensin receptor blocker.

### 3.2 Left ventricular (LV) remodeling

Table 2 presents the changes in volumes and function throughout the first year following MI. There was no significant change in LVEDVi. There was a significant reduction in LVESVi from the second day to the first week (p=0.003) and from one week to two months (p=0.045). From the second month, the LVEDSi was stable. The LVEF increased significantly from two days to one week (p=0.01) and from one week to two months (p=0.001). There was no significant change observed after two months.

**Table 2.** Changes in LV volumes and Ejection Fraction through the first year.

|  | 2 days | 7 days | 2 months | 1 year | p-value |
| --- | --- | --- | --- | --- | --- |
| LVEDVi (mL/m<sup>2</sup>) | 90 ± 14 | 90 ± 16 | 90 ± 17 | 90 ± 22 | 0.98 |
| LVESVi (mL/m<sup>2</sup>) | 48 ± 13 | 45 ± 13 | 44 ± 17 | 44 ± 20 | <0.01 |
| LVEF (%) | 47 (42, 54) | 52 (47, 57) | 52 (50, 60) | 54 (48, 60) | <0.001 |

Volumes are presented as mean ± SD, and LVEF is presented as median (25<sup>th</sup>, 75<sup>th</sup> percentile). P-values <0.05 denotes significant differences between the different time points (One way repeated measure ANOVA for volumes and Friedman test for LVEF ( $\chi^2$ : 30.6, degrees of freedom: 3)). Left ventricular end-diastolic volume index (LVEDVi), Left ventricular end-systolic volume index (LVESVi). Left ventricular ejection fraction (LVEF).

### 3.3 Myocardial infarct (MI) size

The comparison between manual segmentation and the TPM assessment of MI size is presented in Table 3. From the second day to the second month, a significant gradual reduction was observed in both manual absolute (total number of pixels) and relative (percentage of left ventricular myocardial mass) infarct size. There was no significant change in the first week for absolute and relative infarct size derived from the TPM method. Subsequently, a decline in infarct size was observed from day seven to two months, followed by a significant increase from the second month to the first year (p=0.006 and p=0.012 for absolute and relative infarct size, respectively)

**Table 3.** Infarct size by manual segmentation and the Texture-based probability mapping (TPM) method.

|  | Absolute infarct size (no. of pixels in LV) |  |  | Relative infarct size (% of LV mass) |  |  |
| --- | --- | --- | --- | --- | --- | --- |
|  | TPM | Manual | p-value | TPM | Manual | p-value |
| 2 days | 2166 (812, 4631) | 1895 (1274, 3116) | 0.051 | 19.6 (7.1, 34.4) | 16.5 (9.9, 24.3) | 0.047 |
| 7 days | 1870 (770, 3717) | 1956 (681, 2550) | 0.003 | 18.5 (7.1, 27.9) | 16,3 (5.9, 18.3) | 0.003 |
| 2 months | 1459 (338, 2192) | 1259 (564, 1632) | 0.023 | 14.7 (4.0, 20.7) | 11.7 (4.9, 16.2) | 0.002 |
| 1 year | 1552 (865, 3806) | 1242 (761, 2523) | 0.000 | 17.9 (8.5, 32.3) | 14.3 (8.8, 23.1) | 0.001 |

The table compares absolute and relative infarct size between conventional scar assessment by manual segmentation and the TPM method at the four time points. Data are presented as median (25<sup>th</sup>, 75<sup>th</sup> percentile). P-value for Wilcoxon Signed Ranks Test to assess the difference between the manual and the TPM method at the various time points.

### 3.4 Correlations between MI size and biomarkers

NT pro-BNP levels at two months post-PCI were significantly correlated with scar estimation at all four time points assessed by manual segmentation and the TPM method (Table 4). There was a highly significant correlation between absolute TPM infarct size measured at one week with NT pro-BNP at two months post-PCI (r=0.53, p <0.001). Median TnT values with corresponding 25th and 75th: 6.0 (4.2, 10.5) at 11 hours post-PCI. The TPM method exhibited a significant and robust correlation with peak TnT across all four time points (p<0.001), as shown in Table 4.

**Table 4.**
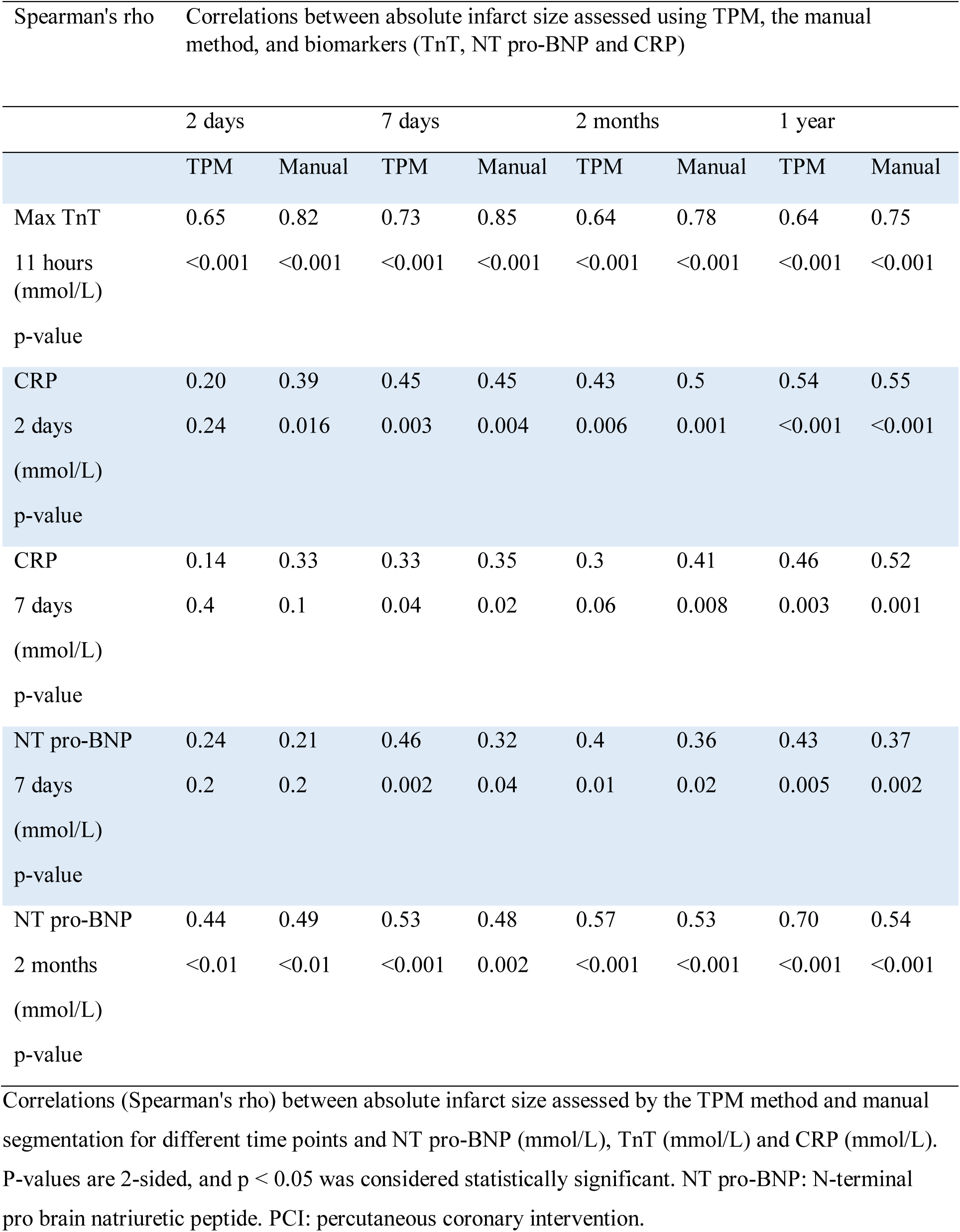
Correlations between infarct Size and biomarkers.

**Table 5.** Biomarkers.

| | Pre PCI | 2 days | 7 days | 2 months | $\chi^2$ | p-value |
| --- | --- | --- | --- | --- | --- | --- |
| TnT<br>(mmol/L) | 0.04 (0.01, 0.13) | 2.46 (1.89, 4.27) | 0.28 (0.10, 0.76) | 0.01 (0.01, 0.01) | 99.7 | < 0.001 |
| CRP<br>(mmol/L) | 3 (1, 8) | 32 (13, 73) | 10 (2,20) | 2 (1, 3) | 94 | < 0.001 |
| NT pro-BNP<br>(mmol/L) | 14 (6, 23.) | 158 (112, 304) | 57 (34, 113) | 36 (21, 78) | 101.8 | < 0.001 |

Data are presented as median (25<sup>th</sup>, 75<sup>th</sup> percentile). P-values <0.05 denotes significant difference using the Friedman test. Degrees of freedom: 2. There was a statistically significant difference in all biomarkers across the four time points.

**Table 6.** Correlations between the TPM method and final scar size and remodeling.

| Spearman's rho | Correlations between TPM-based infarct size (absolute and relative) and remodelling parameters one year post PCI |  |  |  |  |  |
| --- | --- | --- | --- | --- | --- | --- |
|  | 2 days |  | 7 days |  | 2 months |  |
|  | Absolute | Relative | Absolute | Relative | Absolute | Relative |
| LVEDVi | 0.405 | 0.315 | 0.324 | 0.246 | 0.379 | 0.251 |
| p-value | 0.01 | <0.05 | 0.03 | 0.1 | 0.01 | 0.1 |
| LVESVi | 0.503 | 0.428 | 0.466 | 0.395 | 0.553 | 0.453 |
| p-value | 0.001 | 0.006 | 0.002 | 0.01 | <0.001 | 0.003 |
| LVEF | -0.475 | -0.425 | -0.580 | -0.538 | -0.676 | -0.624 |
| p-value | 0.002 | 0.006 | <0.001 | <0.001 | <0.001 | <0.001 |
| Manual MI size (absolute) | 0.70 | 0.65 | 0.75 | 0.66 | 0.73 | 0.6 |
| p-value | 0.001 | 0.001 | 0.001 | 0.001 | 0.001 | 0.001 |

Correlations (Spearman's rho) between TPM-based infarct at various time points and volume and manual infarct size. . P-values are 2-sided, and $p < 0.05$ was considered statistically significant. Left ventricular end-diastolic volume index (LVEDVi), Left ventricular end-systolic volume index (LVESVi).

## 4. Discussion

TPM showed a consistently strong correlation with peak Troponin T levels at all four time points, suggesting that TPM reflects myocardial injury. TPM correlated with NT-pro BNP more than manual LGE assessment of MI size, indicating that TPM may add information on reduced functional capacity in the non-infarcted myocardium. These findings indicate that TPM adds information to traditional LGE analysis on MI healing.

This study suggests that TPM has the potential to give further insight into scar formation and healing. In a previous study, we explored the relationship between MI size assessed by TPM and conventional estimates of scar size in patients with healed MI ^12^. In the subacute phase after a MI, the current study provides additional insights by comparing TPM and conventional LGE-based MI assessment. After an acute MI, imaging results are highly affected by the acute inflammation triggered by ischemia and cardiomyocyte necrosis. The inflammation peaks within 3-5 days after the infarction, followed by a proliferative phase dominated by fibroblast collagen deposition, transforming the MI into a scar during the following weeks. The extent of the inflammation plays a significant role in infarct healing and cardiac remodel^15^. During the following months, the scar matures ^10, 16^. Initially, the myocardial necrosis may increase the size of the infarcted area. However, the subsequent shrinkage of the MI region over the next few weeks is a consequence of infarct healing that may impact the early assessment of MI size. A proliferation phase succeeds the myocardial inflammation within one week after MI, in which the first components of the scar are established.

Consistent with existing literature, the manual MI method demonstrates a decline in absolute and relative infarct size from the first week to two months ^17, 18^. From the second month, manual infarct size shows no significant change. In contrast, TPM shows a significant increase in infarct size from two months to one year, with a strong correlation with BNP. When comparing the yellow and the purple lines in Figure 3, representing the myocardium at two months and one year, respectively, there is a shift toward more fibrosis. These parts of the myocardium are not classified as scarred by the TPM since they have a lower scar probability of less than 0.328. However, these changes can be assessed and visualized using the TPM method. These findings suggest that the rise in TPM-estimated MI size between two months and one year may reflect fibrous changes in the myocardium outside of the MI defined by traditional methods. These changes remain undetected by the manual LGE-based method due to an inadequate increase in pixel signal intensity. However, myocardial fibrosis in the non-infarcted territory may have significant clinical consequences and be associated with reduced myocardial function. The peri-infarct zone is a determinant of ventricular arrhythmia ^19, 20^. TPM-based assessment may, therefore, potentially be used to assess the risk of arrhythmia. However, these potential clinical implications need to be explored by future studies.

TPM is a post-processing method that uses a classifier to recognize textural patterns in LGE-CMR images based on pixel matrices (Fig. 2). The classifier is trained using a reference of LGE-CMR images from patients with old myocardial ischemic scars, allowing the program to detect tissue characteristics related to scarring ^11^. The reference library used in training determines what tissue characteristics the program can detect. TPM may provide a more comprehensive presentation of the infarct region than conventional LGE-CMR images. TPM can present the entire myocardium as a color map according to a probability distribution, while manual segmentation produces a sharp line between scar and normal myocardium. Conventional late enhancement considers the infarcted area uniform, but TPM can detect the inhomogeneous border zone between normal and infarcted myocardium, which may provide a substrate for ventricular tachycardia ^21^. The study of TPM and corresponding LGE-CMR images (Fig. 1) shows that besides the scar, which has the highest probability for injury, some areas distant from the infarcted region have intermediate probabilities. It is unclear whether these intermediate probabilities represent remote fibrosis, and the current study does not allow for a conclusion on this issue. However, prognostic implications of the current findings will be explored in an ongoing study of patients with implanted devices.

**Fig. 2.**
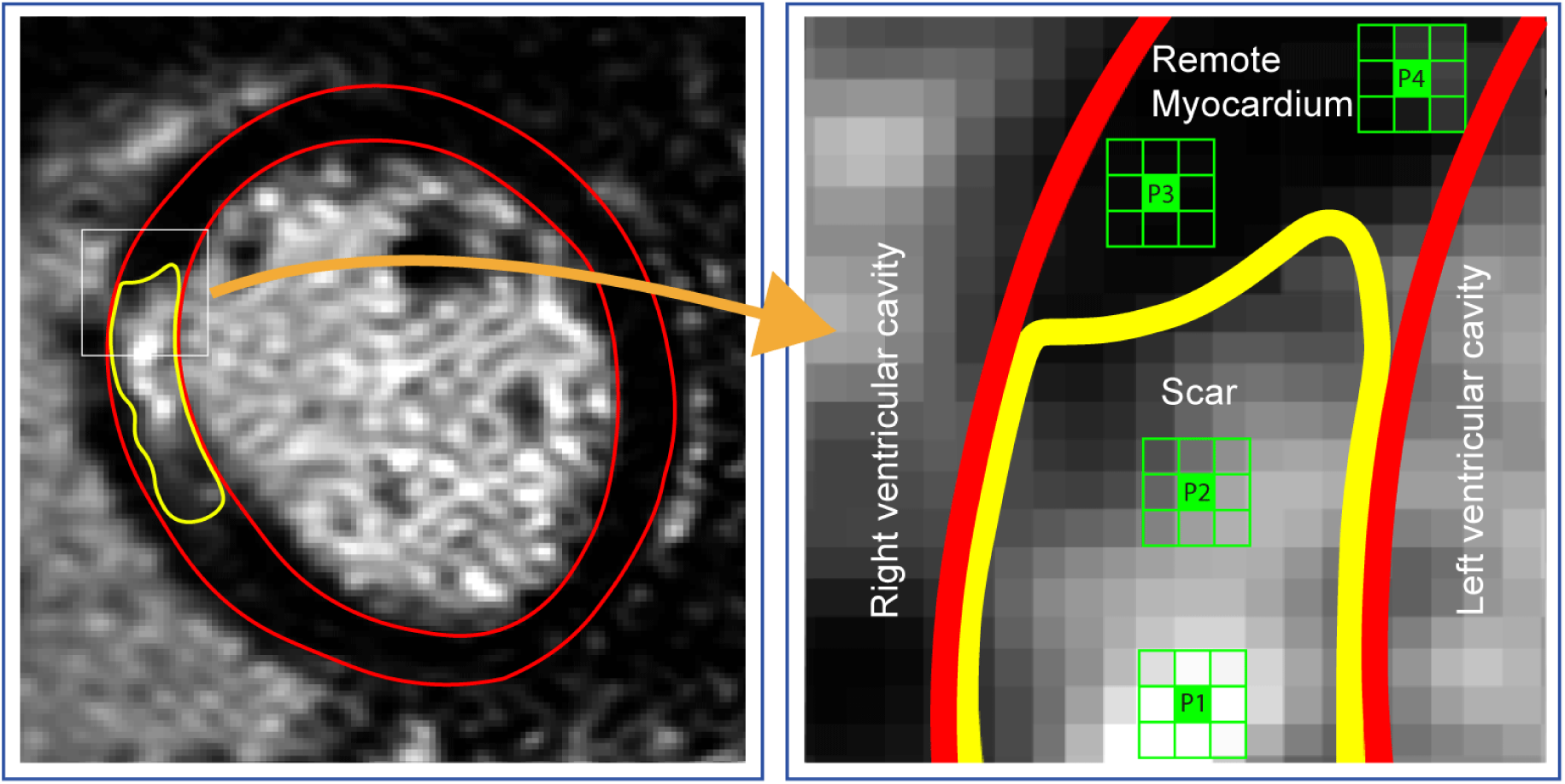
Textural features. The picture to the left shows a Late Gadolinium Enhanced Cardiac Magnetic Resonance cross-sectional image of the left ventricle with the endo-and epicardium delineated manually as the red lines, while the scarred region is delineated in yellow. The picture to the right shows matrices of 3x3 pixels from which the textural features are computed. The pattern within each selected pixel matrix (P1-P4) differs as visualized. These differences demonstrate the heterogeneity within the areas defined by conventional assessment of myocardial infarction.

**Fig. 3.**
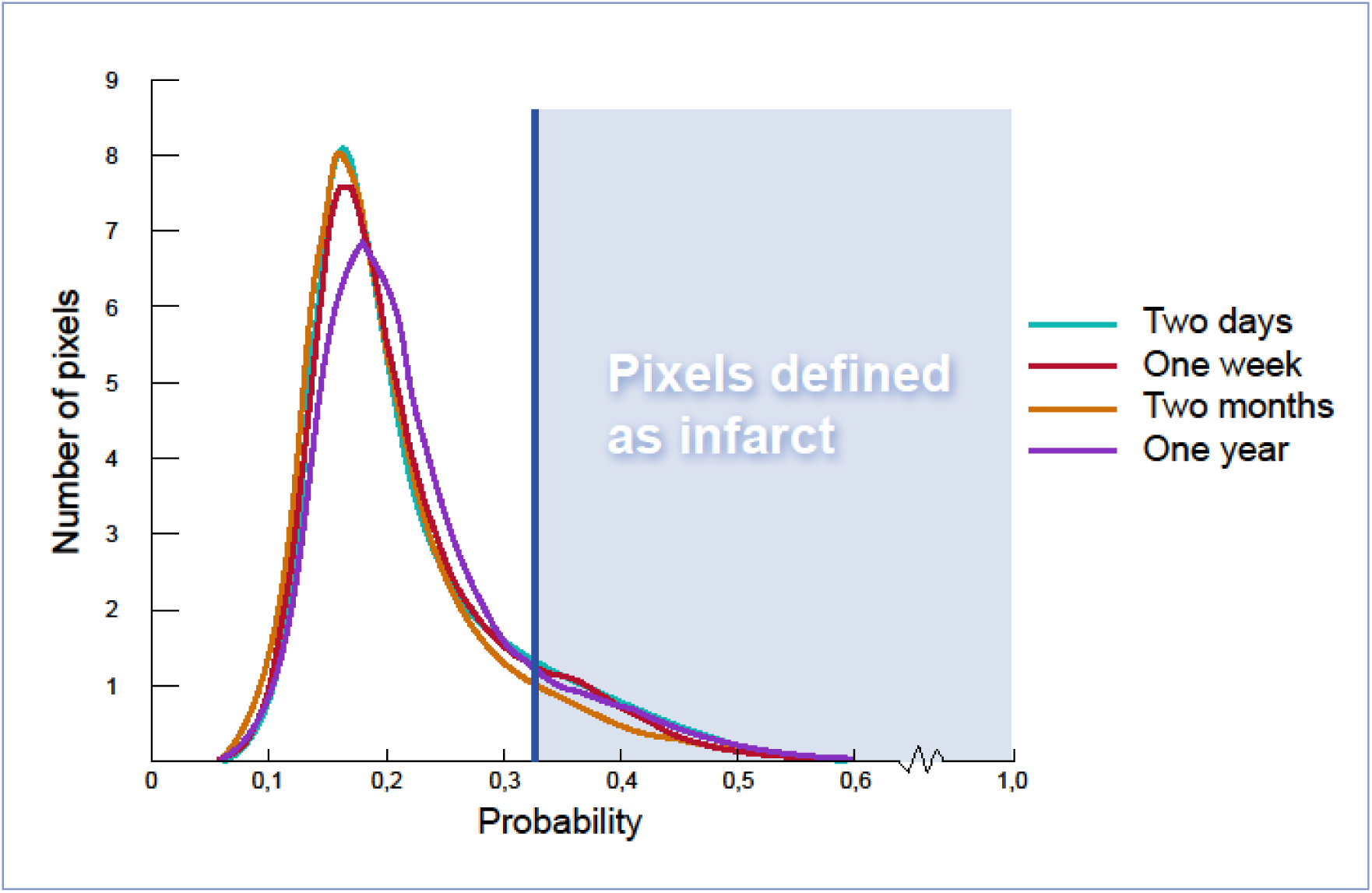
Probability distribution for all pixels at four-time points following acute myocardial infarction. The figure illustrates the total number of pixels (y-axis) with different probability values (x-axis) at each time point of the CMR scan. The lines are made with a density function so that the area under each curve is the same. The blue vertical line is the previously verified cut-off used to determine the probability level (0.328) of pixels associated with myocardial infarction. All pixels with a probability exceeding this level are considered to represent pixels wi areas of myocardial infarction (blue area).

## 5. Study limitations

The study has several potential limitations. First, biological heterogeneity and selection bias may influence the results due to a relatively small sample size. Second, the data used for the present development of the TPM method are based on LGE-CMR images acquired between 2007 and 2009. The use of the current cohort is due to the high LGE image quality and a high-quality homogenous population with successful revascularization of a large, single coronary vessel in patients without any prior MI or established coronary artery disease. All CMR images were acquired without parallel imaging, ensuring high-quality signal density. All images in the current study and the reference dictionary were generated by the same machine (1.5 T Philips Intera R 8.3) using the same methods, contrast agents, and software. These are the same imaging methods used in an ongoing, long-term clinical outcome study assessing the ability of TPM to predict cardiac arrhythmias. Since TPM depends on a reference dictionary, other machines or approaches need to generate a new reference dictionary if they are to use the TPM method. The current study is, therefore, a proof of concept study. If the ongoing clinical study suggests that the TPM method can be used to predict clinical outcomes, the next phase of developing the TPM method needs to generate new dictionaries to test the TPM method in other software and CMR machines.

## 6. Conclusion

Temporal alterations in TPM-based MI size were strongly associated with myocardial injury, inflammation, and increased myocardial work biomarkers. This proof-of-concept study suggests that TPM may add information to traditional LGE-based assessment of acute MI. These findings indicate a potential role for TPM as an additional method for evaluating myocardial injury and fibrosis following acute MI.

## Data Availability

The data that support the findings of this study are available on request from the corresponding author, [V.F]

## Acknowledgements

The authors thank Roald Tungland for his important graphical contributions to the study.

## Declaration of generative AI and AI-assisted technologies in the writing process

The manuscript was written without any use of AI-assisted programs. In the second phase of the preparation of this work, the author used ChatGPT to rephrase some sentences to make them shorter and easier to read. After using this tool/service, the authors reviewed and edited the content as needed and took full responsibility for the publication’s content.

## List of abbreviations

CVD: cardiovascular disease
MI: Myocardial infarction
STEMI: ST-elevation myocardial infarction
MVO: microvascular obstruction
TPM: Texture-based probability mapping
SI: Signal intensity
CMR: Cardiac magnetic resonance imaging
LGE-CMR: late gadolinium-enhanced cardiac magnetic resonance imaging
PCI: percutaneous coronary intervention.
LV: Left ventricular
LVEF: Left ventricular ejection fraction
LVEDVi: Left ventricular end-diastolic volume index
LVESVi: Left ventricular end-systolic volume index
eGFR: estimated glomerular filtration rate
hs-CRP: high-sensitivity C-reactive protein
NT-proBNP: N-terminal pro-B-type natriuretic peptide
TnT: Cardiac troponin-T

